# Deciphering the causal relationship between blood metabolites and Alzheimer’s Disease: a Mendelian Randomization study

**DOI:** 10.1101/2020.04.28.20083253

**Authors:** Jodie Lord, Bradley Jermy, Rebecca Green, Andrew Wong, Jin Xu, Cristina Legido-Quigley, Richard Dobson, Marcus Richards, Petroula Proitsi

## Abstract

There are currently no disease modifying treatments for Alzheimer’s Disease (AD). Epidemiological studies have highlighted blood metabolites as potential biomarkers, but possible confounding and reverse causation prevent causal conclusions. Here, we investigated whether nineteen metabolites previously associated with midlife cognitive function, are on the causal pathway to AD.

Summary statistics from the largest Genome-Wide Association Studies (GWAS) for AD and for metabolites were used to perform bi-directional univariable Mendelian Randomisation (MR). Bayesian model averaging MR (MR-BMA) was additionally performed to address high correlation between metabolites and to identify metabolite combinations which may be on the AD causal pathway.

Univariable MR indicated three Extra-Large High-Density Lipoproteins (XL.HDL) to be on the causal pathway to AD: Free Cholesterol (XL.HDL.FC: *OR*=0.86, 95% *CI=0.78-0.94*), Total Lipids (XL.HDL.L: *OR*=0.88, 95% *CI=0.80-0.97*), and Phospholipids (XL.HDL.PL: *OR*=0.87, 95% *CI=0.81-0.97*); significant at an adjusted threshold of *p*<0.009. MR-BMA corroborated XL.HDL.FC to be amongst the top three causal metabolites, additionally to Total Cholesterol in XL.HDL (XL.HDL.C) and Glycoprotein Acetyls (GP) (posterior probabilities=0.112, 0.113, 0.287 respectively). Both XL.HDL.C and GP also demonstrated suggestive evidence of univariable causal associations (XL.HDL.C:*OR*=0.88, 95% *CI=0.79-0.99*; GP:*OR*=1.2, 95% *CI=1.05-1.38*); significant at the 5% level.

This study offers insight into the causal relationship between metabolites previously demonstrating association with mid-life cognition, and AD. It highlights GP in addition to several XL.HDLs as causal candidates which warrant further investigation. As the pathological changes underpinning AD are thought to develop decades prior to symptom onset, progressing these findings could hold special value in informing future risk reduction strategies.

## 1. Introduction

More than 50 million people worldwide currently live with dementia, and with an aging world population this figure is expected to increase to more than 152 million by 2050 (World Alzheimer Report 2018). The most common dementia type is Alzheimer’s Disease (AD), characterised by impaired everyday function, severe cognitive decline - particularly working, episodic, and declarative memory(1) - and a range of neuropsychiatric symptoms(2). It represents a major source of global morbidity and mortality and poses significant human and economic costs(3).

Disappointingly, AD drug development has proven difficult, with a 99.6% failure rate in the decade of 2002 to 2012, and this rate continues at the same low level today (4). Numerous reasons have been proposed as to why such clinical trials have failed, including incomplete understanding of true causal mechanisms. It is therefore necessary to discover biomarkers that can identify individuals at high risk of developing AD. Moreover, it is important for these to be potentially modifiable so as to offer targets for preventative or therapeutic strategies.

Metabolomics represents one avenue that may give a deeper insight into AD aetiology. Metabolites are small molecules (<1500 atomic mass units) with a role in metabolism(5). As the products of many biological processes, they sit at the end of the systems biology pathway and therefore represent effective intermediate phenotypes to a given disease due to their proximity to the clinical endpoint(6,7). Due to 1) their non-invasive nature of measurement, 2) the fact that they are potentially modifiable through diet and lifestyle, and 3) the ability of many to cross the blood brain barrier, blood metabolites are both practical and valuable markers of biological processes and disease states in dementia(8).

Markers of lipid metabolism have received particular attention in this context, as the impairment of lipid metabolism has been associated with Alzheimer’s disease (5,9-11) and beta-amyloid (Aβ) burden (12,13). Relevant to early intervention, they have also been associated with cognitive performance and brain function during normal ageing(14,15). Recently, using a large British population-based birth cohort, we investigated associations between 233 blood metabolites and both memory and processing speed at 60–64 years of age, as well as changes in these cognitive domains from 60–64 to 69 years old. Associations with several metabolite classes were observed, including fatty acids (FAs), various compositions of high-density lipoproteins (HDLs) and glycoprotein acetyls (GP)(16).

However, it is not yet established whether these metabolites are causally associated with dementia and AD. This study therefore aims to expand our observational findings and assess whether nineteen blood metabolites previously associated with late midlife cognition are causally associated with AD, using several univariable and multivariable Mendelian Randomization (MR) approaches. A deeper knowledge of blood metabolites on the causal pathway for AD will allow for a better understanding of the aetiology of AD and thus facilitate more targeted development and application of future therapeutics.

## 2. Methods

A flow diagram summarising the methodology is detailed in Figure 1.

**Figure 1.**
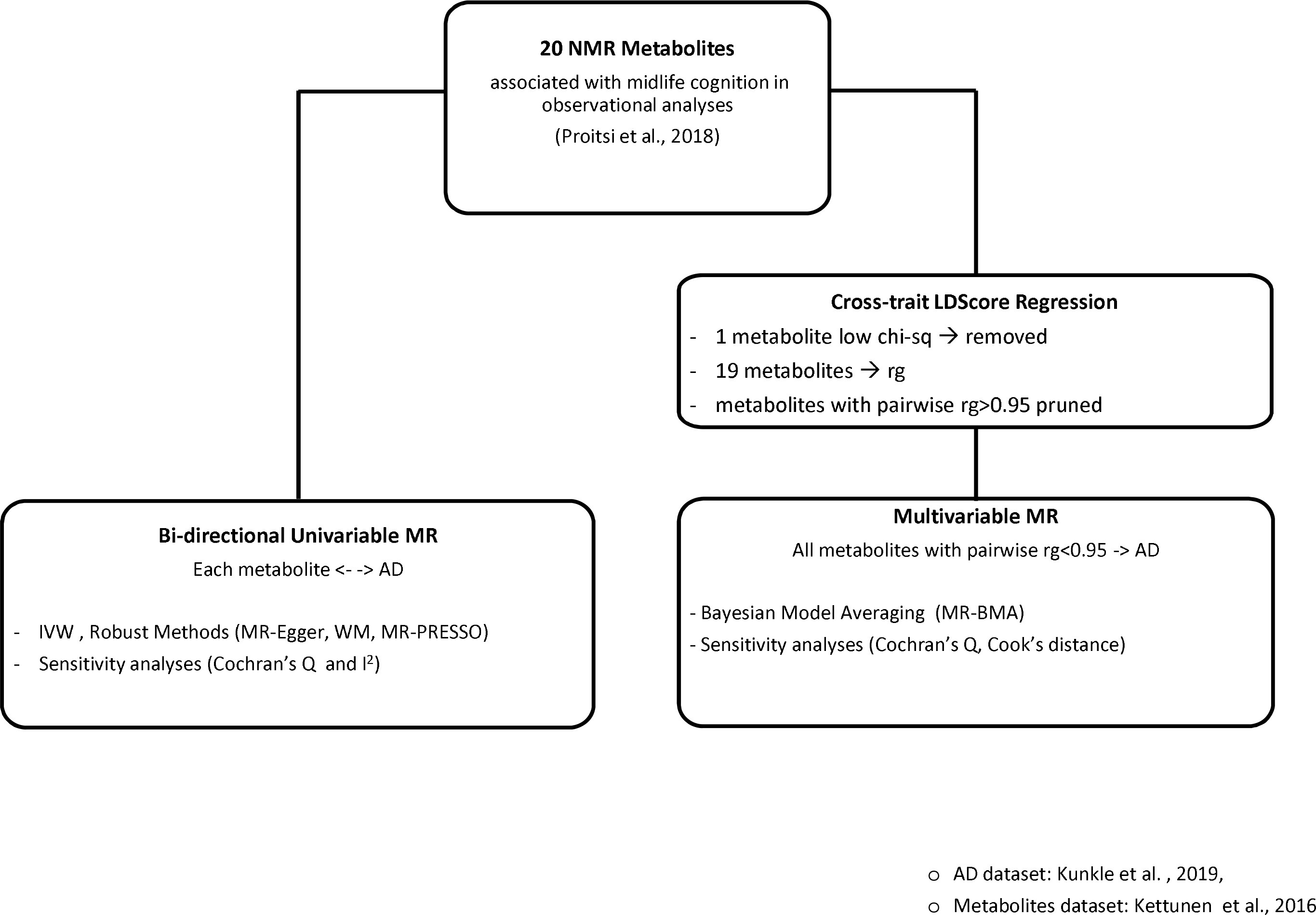
Study design.

**Figure 2.**
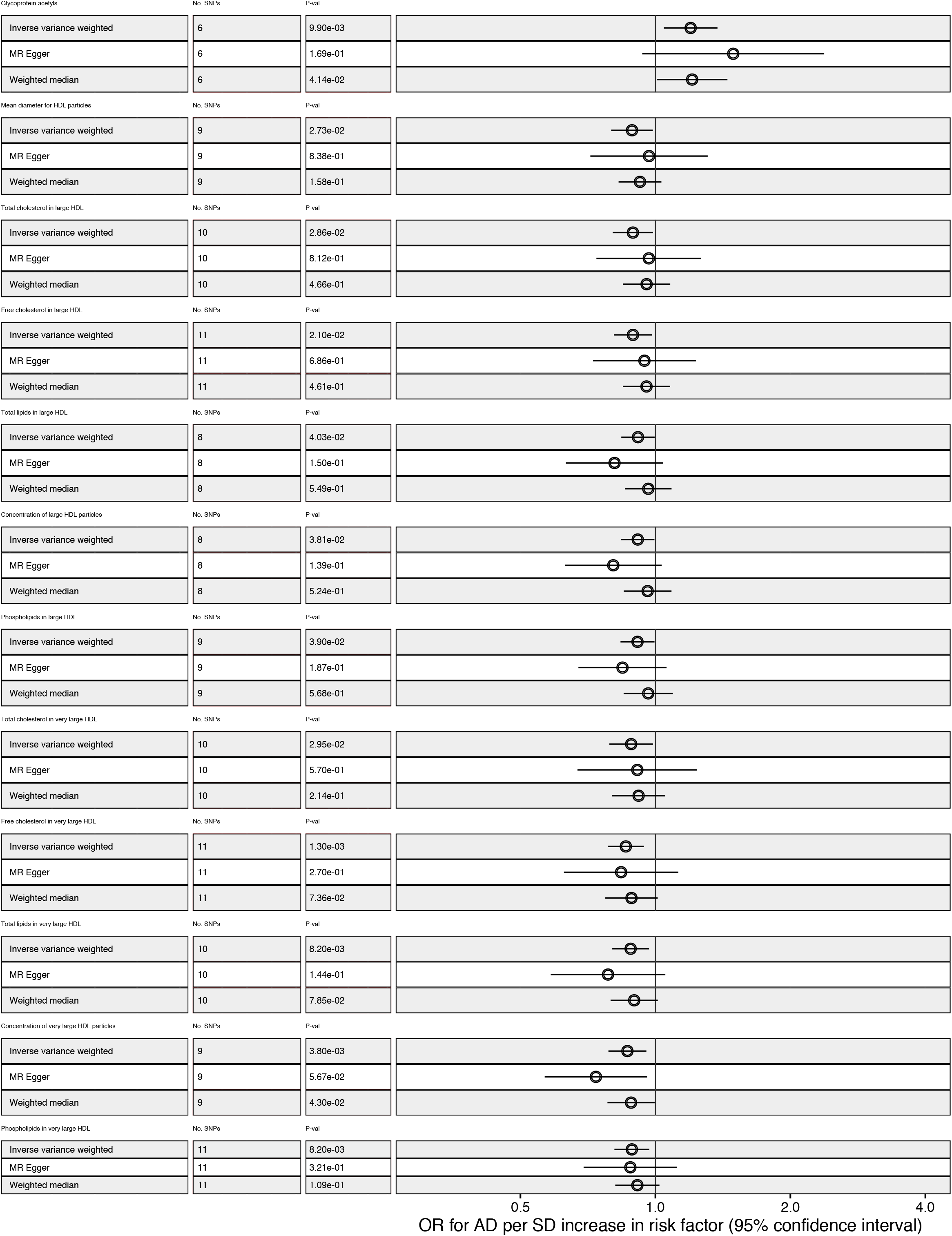
Association of metabolites associated with AD at *p*<0.009 following MR. Results for IVW, MR-Egger and Weighted Median for each metabolite are displayed. Metabolites are sorted based on the beta estimate of the association with AD (IVW).

### 2.1. Metabolite Selection

Metabolites were selected based on our previously published observational study, which investigated associations between blood metabolites and lifetime cognition using data from the MRC National Survey of Health and Development (1946 British birth cohort)(16). Briefly, levels of 233 blood metabolites in 798 participants were measured via nuclear magnetic resonance (NMR) spectroscopy, between ages 60-64(17)(18). At the time of blood-extraction, three domains of cognition were also measured; short-term memory, delayed verbal memory and processing speed(19). These measures were then repeated at age 69 for 663 of the 798 participants(17). Twenty metabolites were significantly associated with at least one measure of cognition after adjusting for multiple testing (*p*<= 0.002) and these were selected for causal investigation within the present study(Supplementary Table S1).

### 2.2. Data sources

For metabolites, summary statistics from the latest and largest metabolite GWAS were used(20) (data: http://computationalmedicine.fi/data#NMR_GWAS). This GWAS investigated the genetic component of 123 blood metabolites on nearly 25,000 individuals using NMR spectroscopy. This platform provides a detailed characterisation of metabolite measures and ratios representing a broad molecular signature of systemic metabolism. Multiple metabolic pathways were covered, including: lipoprotein lipids and lipid sub-classes, FAs and FA compositions, and amino acids and glycolysis precursors. Specific details are described elsewhere(21-23).

Of the twenty metabolites previously associated with cognition, all had at least one single nucleotide polymorphism (SNP) association at genome wide significance (GWS)(*p*<5*10_-8_). However, as only two GWS SNPs were available for Pyruvate, this metabolite was removed due to power concerns, leaving nineteen metabolites for MR. To avoid weak instrument bias, a computed F-statistic of at least 10 was also required for all SNP instruments.

For AD, summary statistics from the latest GWAS of clinically diagnosed late-onset AD (LOAD) by Kunkle and colleagues were utilised(24). This study consisted of three stages; 1) a discovery phase of 63,926 samples, 2) a replication phase of 18,845 samples, and 3) a post replication phase of 11,666 samples. For MR with AD as an outcome, stage 1 summary data were utilised, and for MR with AD as an exposure, stage 1&2 data were employed.

### 2.3. Mendelian Randomisation

#### 2.3.1. Univariable analyses investigating metabolites as causal risk factors for AD

##### SNP Selection

All data extraction, pre-processing, and analyses were performed within R.3.6.1. and using the MRBase package(25). For each metabolite, summary statistics consisting of effect sizes, standard errors and p-values for all GWS SNPs were extracted from each of the GWAS datasets(20). SNPs associated with AD at GWS were excluded due to potential violation of the MR exchangeability assumption(26), which assumes SNP instruments are not associated with confounding risk factors. Any SNPs within the ApoE genomic region (chromosome 19, base-pairs 4500000-4580000) were also excluded for this reason, as ApoE is an established risk factor for traits additional to AD, such as coronary artery disease(27). This resulted in SNP exclusions from large HDL subclasses only (supplementary table S1). Data were harmonised between AD and metabolite datasets, and SNPs with MAF<0.01 were excluded. All GWAS were assumed to be coded on the forward strand, thus no palindromic SNPs were excluded from analyses. However, Additional sensitivity analyses were performed excluding non-inferable palindromic SNPs (MAF>0.40), with metabolite MAFs used to infer AD allele frequencies, due to MAF non-availability within the AD dataset.

##### Mendelian Randomisation analyses

Total causal estimates were computed using inverse variance weighted (IVW) two-sample MR, setting each metabolite as the exposure in turn and AD as the outcome. Briefly, IVW-MR uses a univariable model to regress SNP-instrument associations with an outcome on SNP-instrument associations with an exposure, weighted by the inverse of the variance in SNP-outcome associations(26). To reflect MR’s ‘exclusion restriction assumption’, which states that SNP instrument(s) must only be associated with the outcome via the exposure(26), the IVW intercept is constrained to zero. Results are presented in OR per 1-SD unit to enable a comparison of the magnitude of effect across all exposures.

##### Robust Methods

Two robust methods – MR-egger and weighted median – were utilised to re-estimate casual associations with IVW assumptions relaxed. Briefly, MR-egger re-estimates IVW causal estimates whilst removing the intercept constraint. Large deviations from 0 are taken as evidence of violation to MR’s exclusion restriction and exchangeability assumptions(28); and large discrepancies between egger and IVW estimates are indicative of pleiotropy. Weighted median provided an alternative estimate which remains valid provided 50% of instruments are valid(26). Briefly, causal estimates for each instrument are ordered and weighted by their association strength. The final estimate is then taken as the 50_th_ weighted percentile of the ordered estimate.

##### Sensitivity analyses

Additionally to re-calculating causal estimates with palindromic SNPs excluded (see, SNP selection), several sensitivity analyses were performed to scrutinise the validity of IVW estimates. Influential points were investigated using leave-one-out analyses, and Cochran’s Q was calculated to test for heterogeneity amongst instruments (Q-*p*<0.05 indicating significant heterogeneity). MR Pleiotropy RESidual Sum and Outlier(MR-PRESSO) test was further utilised to identify and correct for potential bias in estimates due to pleiotropy(29). Briefly, this test consists of up to three parts, with 1) the “global test” providing an estimate for the degree of horizontal pleiotropy (significant pleiotropy indicated by *p*<0.05), 2) the “outlier corrected causal estimate” providing a corrected estimate for any significant pleiotropy detected, and 3) the “distortion test” providing an estimate for the degree to which the original and corrected estimates differ (*p*<0.05 indicating a significant difference following corrections for pleiotropy). Tests 2 and 3 are implemented only in cases where *p*<0.05 for global test estimates.

#### 2.3.2. Univariable analyses investigating AD as a causal risk factor for metabolite levels

To explore causality in the opposite direction, AD was set as the exposure with each metabolite in turn set as the outcome. The same analysis pipeline followed as above, testing the association of GWS SNPs from Stages 1&2 of Kunkle et al.(24). Following clumping and the removal of SNPs with MAF<0.01, 24 SNPs were utilised as instrumental variables in causal analyses (Supplementary Table S2).

#### 2.3.3. Multivariable analyses

To account for uncaptured pleiotropy within univariable models and to allow models which capture potential groups of causally associated metabolites, Bayesian model average MR (BMA-MR) was employed(30). This allows for the measurement and selection of multiple, potentially highly correlated exposures in a single model and is particularly well-equipped to scale to high-throughput datasets.

##### Data preparation

BMA-MR adopts a multivariable framework, whereby multiple exposures can be included within the model, provided a) they are each robustly associated with a least one SNP-instrument used within the model, and b) they do not induce multi-collinearity(30). As with univariable models, criterion a) was met through inclusion of only GWS instruments which also had a computed F-statistic of >=10. To meet criterion b), pairwise genetic correlations (*rg*) across metabolites were computed using linkage-disequilibrium score regression (LDSC)(31). Any metabolites with *rg*>0.95 were assumed non-independent and pruned according to the stepwise criteria outlined in Supplementary Information(N1).

##### Bayesian Model Averaging

Following LDSC pruning, nine metabolites were taken forward to MR-BMA (Supplementary Table S3). Instruments were extracted and clumped from each of the metabolite GWASs. Following LD-clumping, removal of ApoE SNPs and removal of a SNP for which a suitable proxy (*R2*>0.8) could not be obtained, 21 instruments remained. As with univariable analyses, all SNPs were assumed to be on the positive strand and sensitivity analyses were performed excluding palindromic SNPs.

Details of the BMA-MR methodology can be found elsewhere(30). Briefly, marginal inclusion probabilities (MIP) for each exposure were computed, representing the posterior probability *(pp)* of metabolite *x* appearing within the true causal model given *z* iterations. Metabolites with highest MIP were interpreted as being the strongest “true causal” candidates of all those provided within the model. A model averaged causal effect (MACE) was also estimated, representing the estimated direct effect of metabolite *x* on outcome *y*, averaged across each *pp*. Here, we set *z* to 10,000, the *pp* to 0.1, and prior variance (σ2) to 0.25.

Q-statistics quantified potential instrument outliers, and Cook’s distance (*Cd*) was used to identify influential points in models with *pp*>0.02. Diagnostic plots were generated to investigate the predicted versus observed associations for each of the top 4 models. Any SNPs with Q-statistic >10 or *Cd*>0.19 (4/total SNP *N)*, were flagged and MR-BMA repeated with the SNP(s) omitted. Metabolite-AD associations remaining after the removal of potential outliers were considered to be more reliably associated with AD.

## 3. Results

### 3.1. Univariable bidirectional MR

For strong evidence of causality, estimates were required to demonstrate association below an adjusted significance threshold of *p*<0.009 (Supplementary Information N2). By this criterion three metabolites retained strong evidence of an IVW causal association with AD: Free Cholesterol in Very Large HDLs (XL.HDL.FC)(*OR*=0.86, *95% CI*=0.78-0.94, *p*=0.001), Total Lipids in Very Large HDLs (XL.HDL.L)(*OR*=0.88, *95% CI*=0.80-0.97, *p*=0.008), and Phospholipids in Very Large HDLs (XL.HDL.PL)(OR=0.89, *95% CI*=0.81-0.97, *p*=0.008). GP also demonstrated evidence of suggestive causal association, with IVW estimates indicating increased odds of AD given higher GP levels (*OR*=1.20 *95% CI*=1.05-1.38); though p-values did not reach adjusted significance (*p*>0.009)(Table 1, Figure 1 and Supplementary Information F1a-Fs).

**Table 1.**
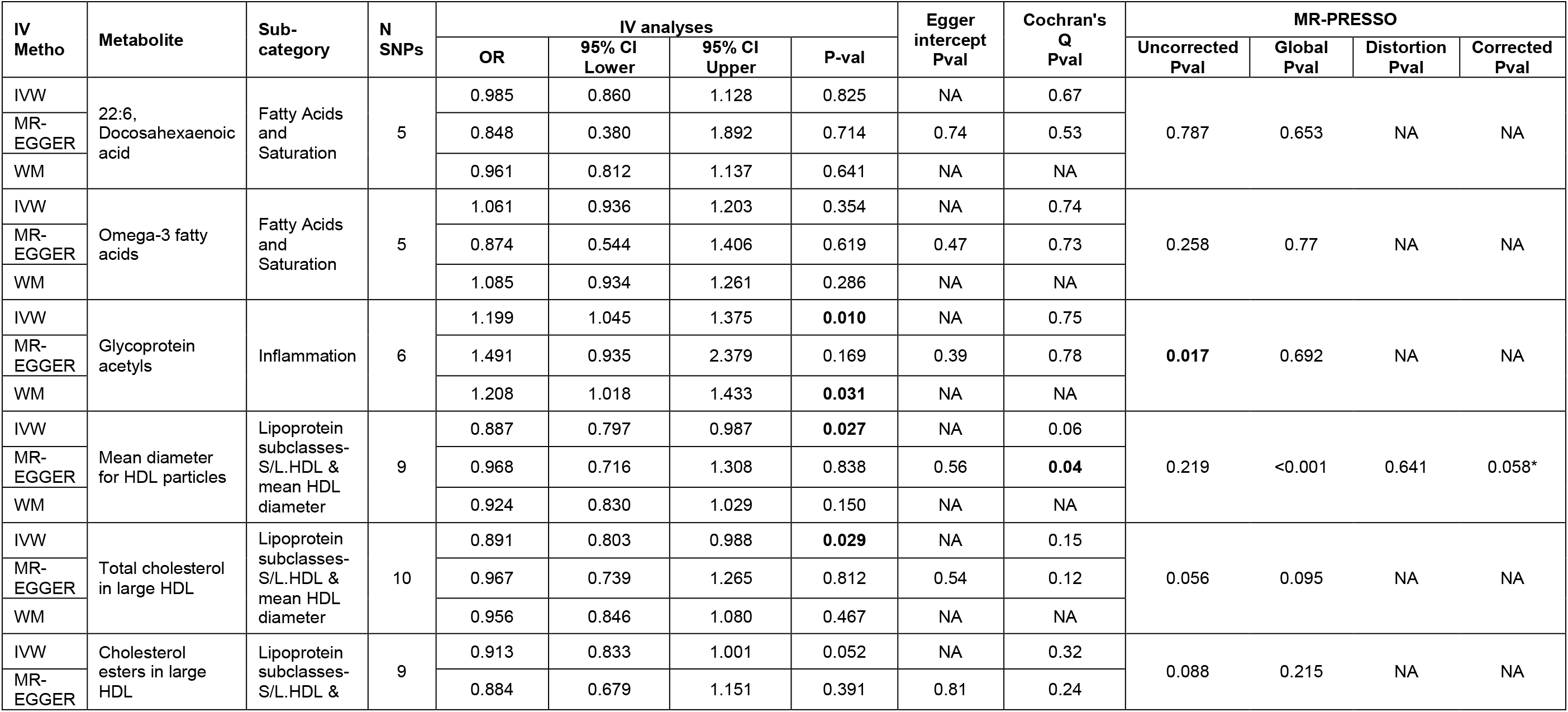
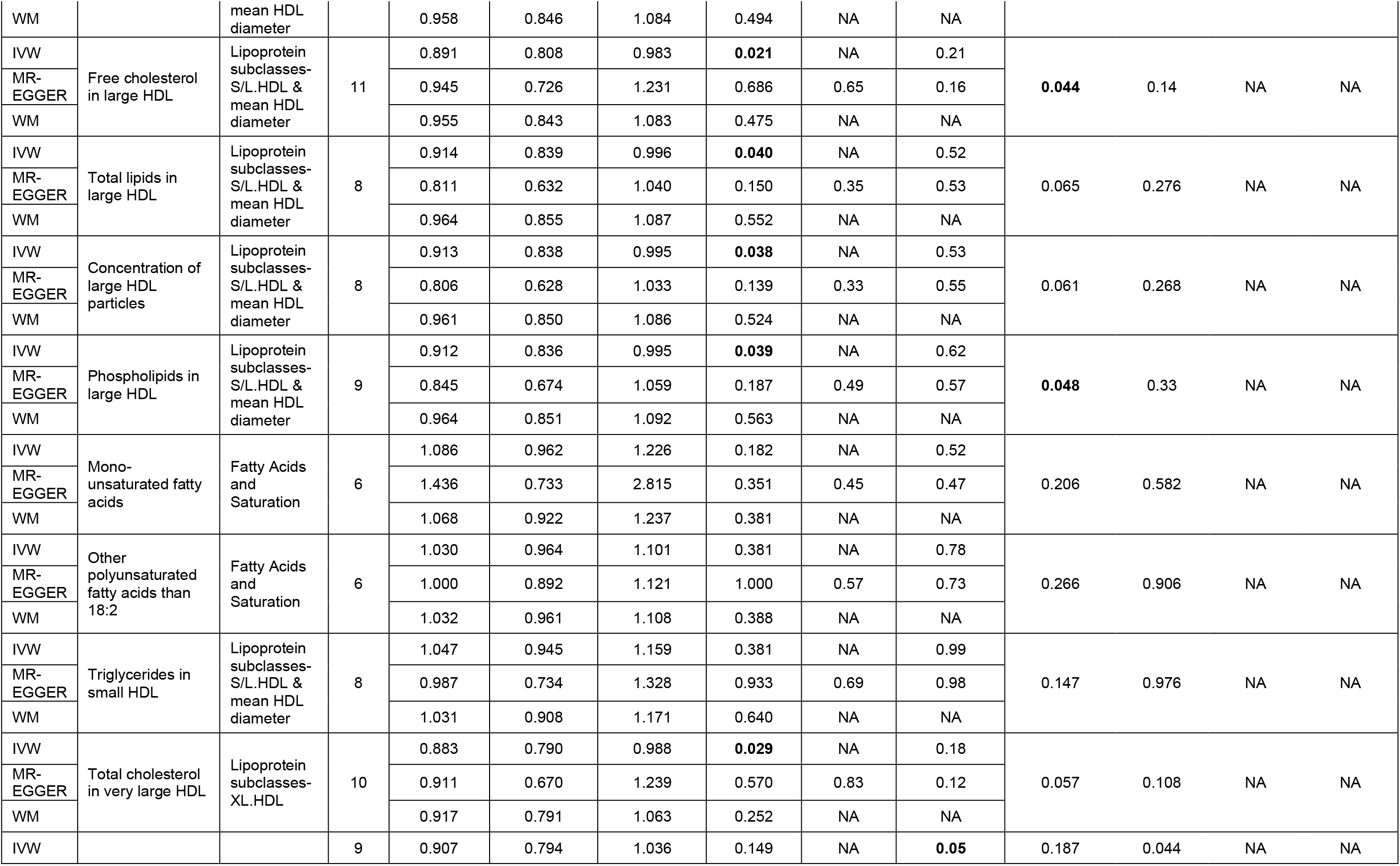
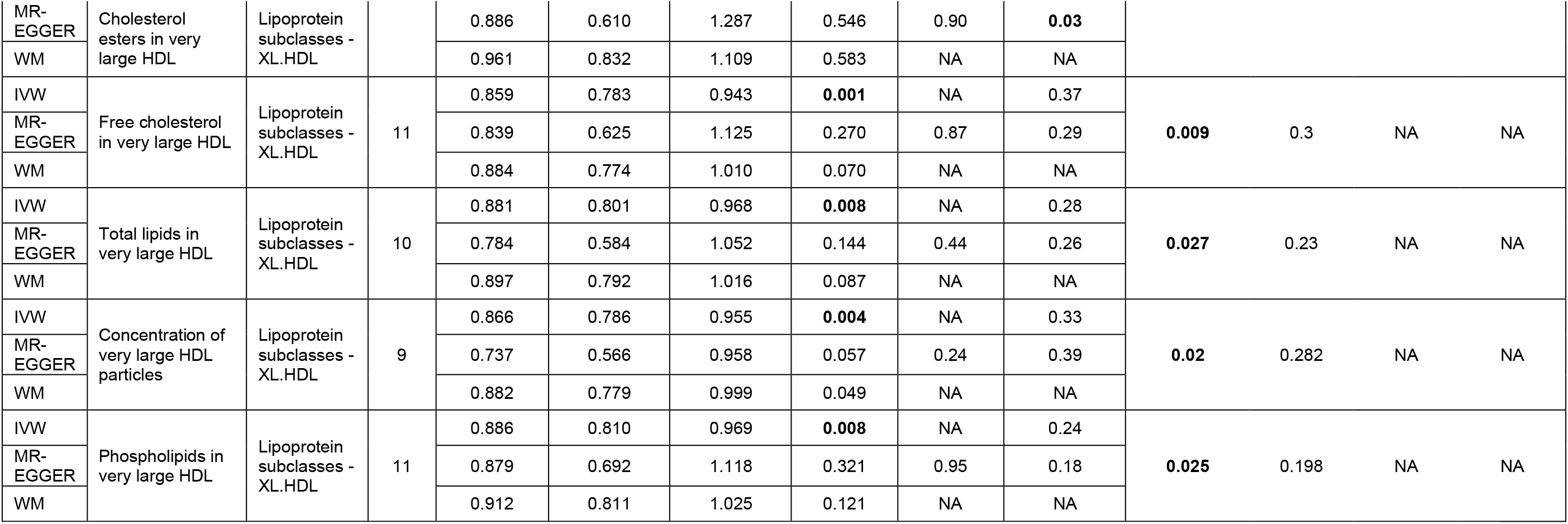
Results of univariable MR analyses investigating the causal association of 19 metabolites with AD. IV-analyses are presented for IVW, MR-Egger and Weighted Median MR. The Egger intercept Pvalue tests for horizontal pleiotropy and Cochrans Q tests for heterogeneity. The corrected P-value of MR-PRESSO test is used when both global and distortion tests were significant.*2 SNPs were highlighted as outliers

Overall, directionality of MR-Egger and Weighted-Median results were in agreement with IVW-MR, though confidence intervals were wider, resulting in a failure to retain significance at the adjusted level. There was no evidence of horizontal pleiotropy as measured with the Egger intercept(Table 1). Funnel plots demonstrated symmetrical distribution of SNP effects around the effect estimate for most tests, suggesting balanced pleiotropy, although this was not the case for metabolites with small SNP *N* (Supplementary Information F2a-F2s).

Leave-one-out indicated two influential SNPs (rs1532085, rs261291) for most HDL sub-fractions, particularly large HDL, and one SNP for GP (rs77303550) (Supplementary Information F3a-F3S). Removal of these SNPs resulted in wider confidence intervals, with only XL-HDL.FC retaining significance at *p*<0.05. Exclusion of non-inferable palindromic SNPs produced almost identical results across IVW, MR-Egger, and weighted median results (Supplementary Table S7).

There was no evidence of a causal relationship in the opposite direction, with AD set as the exposure and each metabolite as the outcome. In addition, there was heterogeneity observed, particularly for very large HDLs, as well evidence of horizontal pleiotropy (Supplementary Table S4, Supplementary Information F4-F7).

### 3.3. Multivariable Bayesian Model Averaging MR

Metabolites ranked by their marginal posterior probabilities (MIP) together with the corresponding average effect are presented in Table 2a, and Table 2b presents the “best” five models based on *pp* rankings. The top risk factor with respect to its MIP was GP (*MIP*=0.465), followed by three XL.HDL particles (XL.HDL.C: *MIP*=0.179; XL.HDL.FC: *MIP*=0.178; XL-HDL-CE *MIP*=0.164). This was corroborated by *pp* rankings, which placed the same four metabolites within the highest ranked causal models, with *pp*s of 0.287, 0.113, 0.112, and 0.102 for GP, XL.HDL.C, XL.HDL.FC, and XL.HDL.CE respectively.

**Table 2.** a) Metabolites ranked by their marginal inclusion probability (MIP) in MR-BMA analyses. b) Best individual metabolite models based on their posterior probability.

| **Metabolite** | **MIP** | **Average Effect** |
| --- | --- | --- |
| GP | 0.465 | 0.088 |
| XL-HDL-C | 0.179 | -0.022 |
| XL-HDL-FC | 0.178 | -0.022 |
| XL-HDL-CE | 0.164 | -0.017 |
| S-HDL-TG | 0.107 | -0.015 |
| L-HDL-C | 0.098 | -0.007 |
| L-HDL-CE | 0.096 | -0.007 |
| DHA | 0.044 | -0.003 |
| PUFA | 0.024 | 0.001 |

b)
| **Metabolite model** | **Posterior Probability** |
| --- | --- |
| GP | 0.287 |
| XL-HDL-C | 0.113 |
| XL-HDL-FC | 0.112 |
| XL-HDL-CE | 0.102 |
| L-HDL-C | 0.050 |
| L-HDL-CE | 0.049 |
| Gp,XL-HDL-C | 0.020 |
| XL-HDL-CE,Gp | 0.019 |
| Gp,S-HDL-TG | 0.019 |

Q-statistics indicated no outliers (Supplementary Information F8a-F8d). The genetic variant with the largest *Cd* was rs1532085, near the *LIPC* gene, which had a *Cd*>0.19 in all three XL-HDL models (XL.HDL.C: *Cd*=1.095; XL.HDL.FC: *Cd*=1.25; XL.HDL.CE: *Cd*=1.168). rs2575876 on the *ABCA1* gene, also demonstrated a high *Cd* in all three XL-HDL models (XL.HDL.C: *Cd* =0.392, XL.HDL.FC: *Cd*=0.247; XL.HDL.CE: *Cd*=0.302), and variant rs247617, near the *CETP* gene had high *Cd* in XL.HDL.C (*Cd*=0.229) and XL.HDL.FC(*Cd*=0.265). Finally, variant rs77303550 on the *TXNL4B* gene, had a high *Cd* in the GP model (*Cd*=0.518), though was <0.19 in all other models (Supplementary Information F9a-Fd). A full overview of Q-statistics and *Cds* for models with *pp*>0.1 are presented in supplementary tables (S5a-S5b).Removal of influential points reduced MIPs, particularly for HDLs, but did not substantially change results (Supplementary Tables S6a-S6b). All MR-BMA results remained consistent when re-ran with non-inferable palindromic SNPs removed (Supplementary Table S8).

## 4. Discussion

This study sought to investigate whether nineteen metabolites previously found to be associated with late mid-life cognition in a British cohort, are on the causal pathway to Alzheimer’s disease (AD). Using summary data from the largest metabolomics and AD GWASs to date, this was interrogated using a combination of both bidirectional univariable and Bayesian multivariable MR. Univariable and multivariable MR highlighted an inverse causal relationship between sub-fractions of extra-large HDL molecules and AD, indicating a protective effect. On the other hand, Glycoprotein Acetyls(GP) – a marker of inflammation – demonstrated evidence of a casual association in the positive direction, indicating that this metabolite may increase AD risk. To our knowledge, this is the first study which uses blood metabolites previously associated with midlife cognition to systematically investigate causal associations with AD.

Sub-fractions of large and extra-large HDLs – in particular their cholesterol content – demonstrated evidence for a causally protective association with AD, supported by both univariable and multivariable results. These results are consistent with our previous observational study which demonstrated positive associations of XL-HDLs and cognition in late mid-life(16). HDLs have also been implicated more widely in age-related cognitive decline and AD(14), with evidence from human studies, animal models, and bioengineered arteries of a cerebrovascular protective effect, which commonly show dysfunction in AD (32). As there is currently no evidence that HDL can enter the brain parenchyma, it is proposed that HDLs circulating in the lumen of cerebral vessels impact brain health through effects on vessel health(32). AD GWASs have further highlighted associations with lipoprotein metabolism, with associations found for SNPs near genes that encode HDL protein components and biogenesis proteins such as *APOE, ABCA1, APOA1 &2, CLU, LCAT and CETPI*(32). Previous MR studies, including ours (33,34) have failed however, to show a causal link between HDL-C levels and AD, potentially due to insufficiently capturing HDL composition complexity. To our knowledge, this study represents the first to provide deeper granularity through inclusion of specific sub-fractions and sizes of HDL.

GPs also demonstrated evidence of positive causal association with AD, particularly in multivariable analyses. This corroborates our previous study, which observed an association between GP and lower cognitive ability in late midlife; consistent with findings from a large independent cohort (14). Additionally, A1-acid glycoprotein has been shown to be a strong predictor of 10-year mortality(35) as well as all-cause mortality in a recent large meta-analysis of >40K individuals(36). Changes in the level of several glycoproteins have also been observed in the hippocampus and inferior parietal lobe in human AD (37). Some of these glycoproteins interact with neurofibrillary tangles, leading to speculation that changes in their glycosylation may be associated with the pathogenesis of this disease (37).

Interestingly, while our previous observational study found the strongest associations to be between fatty acids and late midlife cognition, the present study found no evidence for causal associations between these and AD. This may in part be due to only a low number of instruments available for fatty acids (five SNPs available for both omega-3 and DHA, and six available for MUFA), resulting in a lack of statistical power to detect a causal relationship between these metabolites and AD. Alternatively, this inconsistency could be attributable to the different phenotypes studied (cognition verses AD), with fatty acids potentially being associated with non-AD related cognitive decline, but not AD specifically. Finally, previously observed associations may simply reflect confounding, whereby spurious associations arose as a result of an alternative, non-measured causal component – hence highlighting the importance of methods such as MR for disentangling such scenarios.

Strengths of this study include the use of the largest and most up to date GWASs available for both NMR metabolomics and AD. Moreover, through use of bidirectional MR, relationships were interrogated in both directions as opposed to relying on a-priori (potentially erroneous) assumptions about directionality. Employment of Bayesian model averaging also allowed for correlations between metabolites to be accounted for and for multivariable models of combined metabolites to be proposed. Further, the inclusion of robust and sensitivity analyses across univariable and multivariable models allowed for further interrogation of MR assumptions, ensuring that any notable changes in results could be investigated.

There remain, however, some limitations. First, for several metabolites, less than ten genetic variants were available at genome-wide significance, with two having only five variants available at this level. Whilst steps were taken to ensure individual SNPs did not suffer from weak instrument bias through calculation of per-instrument F-statistics, we cannot exclude the possibility of false negative errors due to insufficient statistical power. Second, due to the absence of available stratified GWA data, the present study was unable to stratify on key variables such as sex – something which our previous observational study indicated may modify many metabolite-cognition associations, and may plausibly too, modify metabolite-AD associations (38). Finally, whilst several IVW causal associations were observed, 95% confidence intervals for robust method estimates were larger, resulting in a loss of significance. Both MR-Egger and weighted median were introduced as a means for re-estimating causal estimates in the presence of potential pleiotropy. Failure of these to detect a causal effect could therefore indicate violation to MR’s core exchangeability and exclusion restriction assumptions. MR-Egger does however, provide an additional test of pleiotropy via its intercept; this indicated no significant pleiotropy across any of our IVW estimates. Moreover, no significant heterogeneity was observed, and consistent directionality for point estimates were maintained across different univariable methodologies. Additionally, BMA-MR – a method able to account for pleiotropy across included exposures – largely corroborated with univariable findings, ranking XL.HDLs and GP as the most likely causal metabolites of those included. Taken together, the weight of evidence supports IVW conclusions, with no indication that wider CIs presented by robust-methods reflect violations to core model assumptions. Instead, it is likely that these reflect low statistical power due to small instrument numbers.

As the pathological changes underpinning AD are thought to develop at least a decade prior to the onset of symptoms, it is important to identify modifiable targets for intervention at an early stage, before AD pathology has caused major irreversible damage. This study suggests that some XL-HDLs associated with late mid-life cognitive ability are potentially causally related to AD, and that GP may too play a role. Progressing these findings could hold special value in informing future risk reduction strategies.

## Data Availability

Data for this work is available within the supplementary material and upon request.

## Acknowledgements

This work was made possible only through generous funding from key funding bodies - PP is funded by Alzheimer’s Research UK and JL is funded by the van Geest endowment fund. This study represents independent research additionally funded by the National Institute for Health Research (NIHR) Biomedical Research Centre at South London and Maudsley NHS Foundation Trust and King’s College London. The views expressed are those of the author(s) and not necessarily those of the NHS, the NIHR or the Department of Health and Social Care.

## Conflict of interests

The authors declare no conflict of interests.

**Supplementary Table S1. Metabolite instruments for Univariable MR analyses**. Instruments included all SNPs that were associated with each metabolite at metabolite GWS.

**Supplementary Table S2. AD instruments for Univariable MR analyses**. Instruments included all SNPs from Kunkle et al., 2019 that were associated with AD at GWS, excluding variants with MAF<0.01 and variants in the APOE gene.

**Supplementary Table S3. Metabolite-Metabolite genetic correlations**. Metabolites were taken forward for Bayesian model averaging MR based on pairwise rg<0.95.

**Supplementary Table S4. Results of univariable MR analyses investigating the causal association of AD with 19 metabolites**.

IV-analyses are presented for IVW, MR-Egger and Weighted Median MR. The Egger intercept Pvalue tests for horizontal pleiotropy and Cochrans Q tests for heterogeneity. The corrected P-value of MR-PRESSO test is used when both global and distortion tests were significant.

**Supplementary Table S5. A) Q statistic and b) Cook’s distance results for the first four MR-BMA models with posterior probabilities >0.1**.

**Supplementary Table S6. a) Metabolites ranked by their marginal inclusion probability (MIP) in MR-BMA analyses and b) Best individual metabolite models based on their posterior probability, after outlier excluding potential outliers rs2575876, rs247617 and rs77303550**.

**Supplementary Table S7. Results of univariable MR analyses investigating the causal association of AD with 19 metabolites after excluding palindromic not inferable SNPs (rs174578, rs609526 and rs2954029)**. IV-analyses are presented for IVW, MR-Egger and Weighted Median MR. The Egger intercept Pvalue tests for horizontal pleiotropy and Cochrans Q tests for heterogeneity. The corrected P-value of MR-PRESSO test is used when both global and distortion tests were significant.

**Supplementary Table S8. a) Metabolites ranked by their marginal inclusion probability (MIP) in MR-BMA analyses and b) Best individual metabolite models based on their posterior probability after excluding palindromic non inferable SNPs (rs2954029)**.

**Supplementary Information N1. Metabolite Pruning in preparation for Bayesian Model Averaging Mendelian Randomization**

**Supplementary Information N2. Multiple test corrections**.

**Supplementary Information F1a-F1s. Scatterplots of the gene-AD versus gene-metabolite associations for each metabolite-AD pair when metabolite is the exposure and AD the outcome following univariable MR analyses**.

Each point in the scatter plot represents an instrumental SNP and different regression lines represents the different MR methods used.

**Supplementary Information F2a-F2s. Funnel plots for each metabolite-AD pair when metabolite is the exposure and AD the outcome following univariable MR analyses**.

**Supplementary Information F3a-F3s. Leave-one-out (LOO) plots for each metabolite-AD pair when metabolite is the exposure and AD the outcome following univariable MR analyses**.

**Supplementary Information F4. Results of univariable MR analyses investigating the causal association of AD with metabolites**.

Results for IVW, MR-Egger and Weighted Median for each metabolite are displayed. Metabolites are sorted based on the beta estimate of the association with AD (IVW).

**Supplementary Information F5a-F5s. Scatterplots of the gene-metabolite versus gene-AD associations for each AD-metabolite pair when AD is the exposure and each metabolite the outcome following univariable MR analyses**. Each point in the scatter plot represents an instrumental SNP and different regression lines represents the different MR methods used.

**Supplementary Information F6a-F6s. Funnel plots for each AD-metabolite pair when AD is the exposure and each metabolite the outcome following univariable MR analyses**.

**Supplementary Information F7a-F7s. Leave-one-out (LOO) plots for each AD-metabolite pair when AD is the exposure and each metabolite the outcome following univariable MR analyses**.

**Supplementary Information F8a-F8d. Diagnostic plots for outliers for the top four models with posterior probabilities >0.1**.

Predicted associations (x-axis) are plotted against observed associations (y-axis) for AD. Any genetic variant with a qvalue>10 is marked with the name of the variant.

**Supplementary Information F9a-F9d. Diagnostic plots for influential genetic variants for the top four models with posterior probabilities >0.1**.

Predicted associations (x-axis) are plotted against observed associations (y-axis) for AD. Any genetic variant with a Cook’s distance >0.19 (4/21) is marked with the name of the variant.

